# Association of *NRAMP1*-INT4 gene with Susceptibility to Tuberculosis in the Sudanese Population: A Case-Control Study

**DOI:** 10.1101/2021.03.15.21253579

**Authors:** Sara Gamal Gubara Mohamed, Arwa Elaagip, Maryam Atif Salaheldin, Abeer Babiker Idris, Taha Hussein Musa, Fayad Osman Mohammed, Layla Ahmed Mohammed, Hassan Hussein Musa

**Affiliations:** Department of Medical Microbiology, Faculty of Medical Laboratory Sciences, University of Khartoum, Khartoum, Sudan; Department of Parasitology and Medical Entomology, Faculty of Medical Laboratory Sciences, University of Khartoum, Khartoum, Sudan; Biomedical Research Institute, Darfur University College, Darfur, Sudan; Institute of Endemic Diseases, University of Khartoum, Khartoum, Sudan

**Keywords:** *M. tuberculosis*, MDR, *NRAMP1*, SNP, Sudan

## Abstract

**Background:** Sudan is a Sub-Saharan African country with a high prevalence rate of Tuberculosis. Natural Resistant Associated Macrophage Protein 1 (NRAMP1) plays a potential role in the development of immunity against TB, and it has a critical role in disease resistance. The aim of the study was to evaluate the association of *NRAMP1* polymorphism at intron4 (INT4) region with susceptibility to TB infection.

**Methods:** Demographic, clinical and microbiological data were collected from 150 participants and investigated using designed questionnaire. The genotyping of *NRAMP1*-INT4 polymorphism was performed in 60 TB-infected patients and 50 healthy control using Polymerase chain reaction and restriction fragment-length polymorphism method (PCR-RFLP).

**Results:** Among cases (60%) were males, only (3.3%) were vaccinated. The most reported risk factors were tobacco smoking (17%), diabetes (10%), alcohol consumption (2%) and corticosteroid therapy intakes (1%). Pulmonary TB was detected in 67% of the patients, 24% had pulmonary/MDR and 9% had extra-pulmonary TB. The frequency of wild G allele was significantly higher in cases compared with healthy control subjects (P-value <0.0001). Also, a significant association was observed between the heterozygosity for *NRAMP1*-INT4 variant and resistance to TB infection (P-value 0.001, OR= 4.83, 95%CI 1.96∼11.88). Homozygotes mutant INT4 (C/C) genotype was not detected in both cases and controls.

**Conclusions:** the *NRAMP1*-INT4 polymorphism may serve as marker of unidentified genetic factors that may play a critical role in host immunity to TB in the Sudanese population. Further studies with large sample size are recommended to determine population-specific genetic associations with TB susceptibility in order to guide TB therapy and prophylaxis in a population-specific manner.

## Background

Tuberculosis (TB) is a chronic bacterial disease caused by *Mycobacterium tuberculosis* (*M*.*tb)* which found normally in the environment ^(1)^. Transmission of infection occurs via inhalation of droplet nuclei from infected patients during coughing, sneezing or talking and mainly infect lungs ^(2)^. It’s accounting 2.5% of global burden diseases with more than 90% of total cases occurring in developing countries ^(3, 4)^. In the last report of the World Health Organization (WHO), they estimate that currently about 10 million people developed TB and 1.4 million died in 2019 in which the highest burden is in adult men, who accounted for 56% of all TB cases, compare to adult women who accounted for 32% and children for 12%. Among all TB cases, 8.2% were among people living with HIV ^(5)^. African countries contribute 31% of new cases mostly concentrated in Nigeria and Ethiopia ^(6, 7)^. Among all cases infected with TB, only 5-10% will develop active disease during their lifetime, this influenced by several risk factors which directly related to the host such as socioeconomic, behavioral and physiological factors or indirectly associated with susceptibility to infection such as genetic host factors ^(3, 8-10)^. It’s indicated that, 75% of the patients have more severe form of disease which is exclusive pulmonary involvement (PTB) ^(11)^. Extra-pulmonary TB (EPTB) is the occurrence of TB at sites other than lungs. Globally, the prevalence of EPTB among new and relapse TB cases in 2016 was 15%, the lowest was recorded in western Pacific Region (8%) while the highest was recorded in the Eastern Mediterranean (24%) in which 16% was in African Region ^(11)^. Several high-income countries have reported an increased frequency of EPTB over time ^(11, 12)^.

Sudan is Sub-Saharan African country with population approximately 41 million, ranked at 167 out of 188 countries in the Human Development Index (HDI) with 4.8 million people needing humanitarian assistance because of conflict, environmental and food insecurity factors ^(13, 14)^. It accounts for 14.6% of total TB burden in the Eastern Mediterranean region, this percent ranks Sudan among the high prevalence countries ^(15)^. In 2017, the estimated TB incident rate in Sudan was 77 per 100.000 individual, with number of 20.438 new and relapse TB cases including HIV co-infected ^(16)^. Tuberculosis care and treatment is provided by National Tuberculosis Control Program under the auspices of the Ministry of Health and by several non-governmental organizations (NGOs) who provide care to displaced persons, including those who living in refugee camps ^(17)^. There were several National referral centers for tuberculosis diagnosis and management in Sudan. Abu Anja Specialized TB hospital is one of those centers located in Omdurman, Khartoum state ^(18)^. The diagnostic methods in most of TB centers depends on X-ray imaging, staining by Ziehl-Neelsen Stain and molecular technique of GeneXpert which have been available in 13 states laboratories. As the time of this study, the National reference laboratory located in Khartoum state was the only laboratory with the capacity to perform Mycobacterium drug susceptibility testing (DST) ^(19)^.

Several genes were associated with inflammatory and cellular immune response, triggering a protective action against tuberculosis infection ^(20)^. Natural Resistant Associated Macrophage Protein 1 (NRAMP1) is a candidate gene, that have been reported in many studies of its critical role in disease resistance in humans, mice’s, cattle’s and pigs ^(8, 21-25)^. The human homologues of *NRAMP1* gene has been cloned and mapped to chromosome 2q35 ^(26)^. It has been thought to be a strong candidate gene for human tuberculosis susceptibility, and several studies were performed to evaluate relationships between *NRAMP1* and tuberculosis ^(26, 27)^. Eleven variants were identified in gene locus as single nucleotide polymorphism (SNP) in certain regions, four of them were found to have a significant genetic association with active tuberculosis infection including; 3′UTR (untranslated region), D543N (G/A: Asparagine/Aspartic acid), 5′ promoter (GT)n and INT4 (Intron4: 469+14G/C) ^(28-30)^.

Studying the host genetic profile using molecular approaches may help in the understanding of clinical diversity; and improve therapeutics, vaccination global control of tuberculosis infection ^(31)^. Many studies have found an association between *NRAMP1* gene polymorphisms and the progression of TB to severe forms of pulmonary tuberculosis, rather than susceptibility to *M. tuberculosis* infection ^(32)^. However, to our knowledge, this association has not been studied in the Sudanese population. Thus, the aim of this study was to investigate whether *NRAMP1* variant at INT4 region was associated with susceptibility and/or severity of tuberculosis among Sudanese patients using PCR-RFLP method.

## Methods

### Study setting and study population

The study involved hundred patients with active pulmonary and extra-pulmonary infection, referred to Abu Anja Specialized Tuberculosis Hospital. All patients were ethnically mixed and came from different states in Sudan. Patients with active PTB and PTB/MDR were selected based on abnormal chest X-ray confirmed by results of conventional bacteriological examination including; smear positive for acid fast bacilli (ZN stain) and GeneXpert result for MDR-TB. The EPTB were selected based on bacteriological examination and magnetic resonance imaging (MRI) picture for the extra-pulmonary located infection. For genetic study, fifty control persons were recruited from the Blood Bank Center of Soba Teaching Hospital and selected based on their healthy appearance and no history of TB infection. All subjects in the study were interviewed for collection of data using structured well designed questionnaire and any participant who had chronic illness or one of the risk factors which can effect on the result outcome including those with HIV positive or known to have any autoimmune or chronic inflammatory diseases were excluded. Informed consent was taken from all participants before collecting the blood samples. Approval and clearance for the study was obtained from Abu-Anja Specialized TB Hospital and the research board of the Faculty of Medical Laboratory Sciences, University of Khartoum. 2ml of venous blood collected in Ethylene-Diaminetetraacetic acid (EDTA) vacutainer were taken to the university laboratory and stored at −20°C for Deoxyribo Nucleic Acid (DNA) extraction for detection of gene polymorphism.

### *NRAMP1* genotype analysis

Genomic DNA was extracted from whole blood samples using two types of DNA extraction kits; G-spin^™^Total DNA Extraction Kit and QIAamp DNA Blood Mini Kit in accordance with the manufacturer’s recommendation and quality was evaluated using 1% agarose gel electrophoresis to detect the integrity and sharp bands of DNA. The polymorphic variant (INT4 region) in *NRAMP1* gene was studied by PCR-RFLP technique, using sense primer (5′-GTCTGCCATCTCTACTACCCTAAGGTG-3′) and antisense primer (5′-CATGTCCCTCTAGGTATGTGCTATCAG-3′). Product size was 510bp. Primer quality was checked by OLIGO^™^ software before use. The iNtRON’s Maxime PCR PreMix kit were used for PCR, the tube contains ready mixtures of i-Taq^™^ DNA polymerase, dNTPs mixture, reaction buffer and gel loading buffer mixed with de-ionized water, genomic DNA and primers. PCR amplifying procedure was as follows: 2 min at 95^°^C, 30 cycles of 30 sec at 95^°^C, 30 sec at 65^°^C, 60 sec at 72^°^C then 5 min at 72^°^C, performed on SENSQUEST lab cycler automated thermal cycler. Product was checked on 1% agarose gel electrophoresis in SCIE-PLAN (vision machine) and visualize by digital image analysis system for confirmation of the length and integrity of bands. The INT4 region was analyzed for the presence of SNP using PCR product digested with the specific restrictive endonuclease enzyme *Apa* I (New England Biolabs) reported by Taype et al., 2006 ^(33)^.

### Statistical Analysis

Data were analyzed using statistical package for social science (SPSS statistics 25). We used the unpaired student’s t-test (for continuous numerical data) and Chi-square (X^2^) test (for categorical data) ^(34)^. Odds Ratio (OR) and 95% confidence interval (95%CI) were calculated to quantitatively assess the degree of association between single nucleotide polymorphism (SNP) and tuberculosis infection and between patients and control group. The differences considered statistically significant as P-value <0.05 ^(22)^.

## Results

### Baseline characteristics of the study population

As illustrated in Table 1., TB infection were mostly reported in males (60%) with average age of 32.8±11.83 years. Collective data were taken from each patient considered the risk factors for developing tuberculosis infection includes; BCG vaccination, tobacco smoking, diabetes, alcohol consumption and corticosteroid therapy. BCG immunization was determined by the presence of a typical scar on either upper arm at the time of blood collection and the result showed that only 3.3% of the patients have vaccination scar. The most reported risk factors among tuberculosis patients were tobacco smoking (17%). It was also recorded that 10% of patients have diabetes mellitus, 2% consumed alcohol and 1% received corticosteroid therapy. A total of 67% patients had pulmonary TB, 24% diagnosed as pulmonary/MDR-TB and 9% extra-pulmonary TB (three with extra-thoracic lymphatic, four with bone& joints and two with stomach TB). Most of TB infected patients were diagnosed by smear positive ZN stain and chest X-ray findings (83.4%). All suspected drug resistant MDR cases were diagnosed additionally with GeneXpert (50%) and the other extra-pulmonary TB suspected patients were diagnosed by MRI (3.3%). Fifty-eight percent of patients whom isolated were classed as new patients start receiving first line anti-tuberculosis drugs in the same month. Of the reminder 20% have previously complete the first line drugs classed as MDR-TB patients and started treatment with the second line anti-tuberculosis regimens, 17% were classed as new tuberculosis patients who did not enrolled in the treatment system yet and 5% had experiences interrupted treatment.

**Table 1.** Demographic and clinical characteristics of the patients with tuberculosis infection (n=100)

| Variable | Frequency (%) |
| --- | --- |
| **Gender** |  |
| Male | 60 (60%) |
| Female | 40 (40%) |
| **Age group** |  |
| ≤ 30 | 51 (51%) |
| 31-40 | 26 (26%) |
| 41-50 | 16 (16%) |
| ≥ 50 | 7 (7%) |
| **Marital status** |  |
| Single | 58 (58%) |
| Married | 42 (42%) |
| **Residence** |  |
| Khartoum | 22 (22%) |
| Omdurman | 60 (60%) |
| Gazira | 13 (13%) |
| Nyala | 1 (1%) |
| White Nile | 1 (1%) |
| Kordufan | 2 (2%) |
| Sinnar | 1 (1%) |
| **Risk factors** |  |
| Smoking | 17 (17%) |
| Diabetes | 10 (10%) |
| Alcohol abuse | 2 (2%) |
| Corticosteroids therapy | 1 (1%) |
| **TB forms** |  |
| Pulmonary TB | 67 (67%) |
| Pulmonary/MDR | 24 (24%) |
| Extra-pulmonary | 9 (9%) |

### Association of *NRAMP1*-INT4 with tuberculosis

A total of 60 patients were included for the analysis of polymorphic change in the INT4 loci of *NRAMP1* and compared with 50 healthy individuals belonging to the same geographical area with no history of previous TB infection. Among the selected cases, 36 were males and 24 were females. The distribution of TB forms was; 50% pulmonary TB, 36.7% pulmonary/MDR and 13.3% were extra-pulmonary TB. The homozygous INT4 (G/G) genotype was identified in 51 (85%) cases with tuberculosis and 27 (54%) healthy control subjects, while the heterozygous INT4 (G/C) was found in 9 (15%) cases and 23 (46%) healthy controls (P-value 0.001, OR= 4.83, 95%CI 1.96∼11.88) (Figure 1). The homozygous INT4 (C/C) genotype was not detected in neither cases nor healthy controls. The frequency of G allele was significantly higher in cases compared with healthy control subjects (P-value <0.0001). Also, a significant association was observed between the heterozygosity for *NRAMP1*-INT4 variant and resistance to TB infection (P-value 0.001, OR= 4.83, 95%CI 1.96∼11.88) (Table 2).

**Figure 1.**
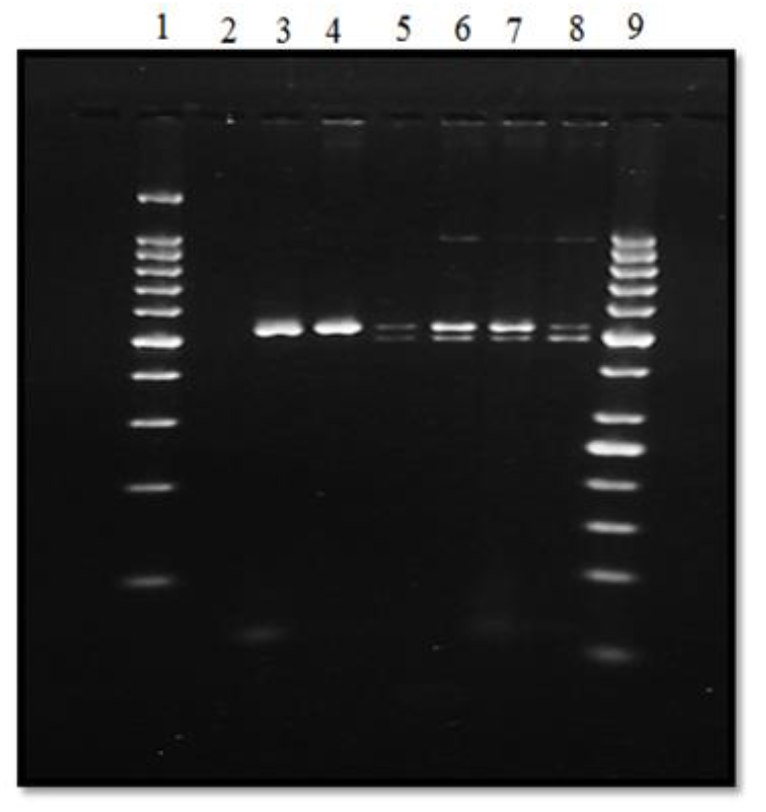
Agarose gel electrophoresis of PCR-RFLP products of cases and healthy controls. 100bp DNA size marker= 1; 50bp DNA size marker=9; Negative control= 2; PCR product= 3; G/G= 4; G/C genotype= 5,6,7,8.

**Table 2.** Genotypes and Allele Frequency of INT4 Polymorphism in Patients and Healthy Controls

| Polymorphisms | Patient with TB n(%) | Healthy controls n(%) | P-value | Odds ratio (95%CI*) |
| --- | --- | --- | --- | --- |
| **Genotype** |  |  |  |  |
| **G/G** | 51 (85) | 27 (54) | 0.001 | 1 |
| **G/C** | 9 (15) | 23 (46) |  | 4.83 |
| **C/C** | 0 | 0 |  | (1.96~11.88) |
| **Allelic frequency** |  |  |  |  |
| **G** | 0.93 | 0.23 | <0.0001 |  |
| **C** | 0.08 | 0.77 |  |  |

\*CI= Confidence Interval

The heterozygous G/C variants were identified in 23.3% pulmonary TB, 4.5% pulmonary/MDR and 14.3% extra-pulmonary TB patients. Significant differences were found between patients with pulmonary TB, pulmonary/MDR and healthy controls. However, there were no significant differences or statistical association between G/C genotype and extra-pulmonary TB (P-value 0.074; OR G/C (95%CI) = 0.167) (Table 3).

**Table 3.** Association between *NRAMP1*-INT4 polymorphism and clinical forms of tuberculosis

| Genotype | Control<br>(n=50) | Pulmonary TB<br>(n=30) | OR<br>(95% CI) | Pulmonary/MDR<br>(n=22) | OR<br>(95% CI) | Extra-pulmonary TB<br>(n=8) | OR<br>(95% CI) |
| --- | --- | --- | --- | --- | --- | --- | --- |
| G/G | 27 (54%) | 23 (76.7%) | 1 | 21 (95.5%) | 1 | 7 (87.5%) |  |
| G/C | 23 (46%) | 7 (23.3%) | 0.357<br>(0.129-0.983) | 1 (4.5%) | 0.055<br>(0.007-0.448) | 1 (14.3%) | 0.167<br>(0.19-1.465) |
| P-value |  | 0.042 |  | 0.001 |  | 0.074 |  |
| X<sup>2</sup> |  | 4.110 |  | 11.814 |  | 3.190 |  |

## Discussion

Genetic variants in the *NRAMP1* gene were shown to associate not only with increased susceptibility to TB infection but also with an increased tendency to develop severe outcome ^(35-37)^. However, different studies have been published analyzing the contribution of *NRAMP1* gene INT4 polymorphism to TB susceptibility and/or severity with conflicting results explained, in part, by ethnic differences. A positive association was observed in West African, American, Asian, Greek and Mestizo Peruvian populations ^(8, 33, 34, 38)^, but there was no association in Korean, Moroccan, Iranian, Thai, Taiwanese, Turkish and Indonesian populations ^(23, 39-44)^. Interestingly, in the present study, we found an association between heterozygosity for *NRAMP1*-INT4 variant and resistance to TB infection (P-value 0.001, OR= 4.83, 95%CI 1.96∼11.88). This finding is, partially, in agreement with studies conducted in Cambodian and Japanese populations in which an association between heterozygosity for polymorphic *NRAMP1* variants (D543N and 3′UTR) and resistance to TB were reported ^(1, 45)^. The discrepancy between our finding and others could be explained by understanding whether polymeric *NRAMP1*-INT4 variant itself confers a truly altered susceptibility to TB infection and is functional or the associated allele is in linkage disequilibrium with an unknown disease susceptibility allele. In this regard, Søborg *et al*. suggested that the *NRAMP1* loci (INT4, G543A and 3′UTR) might be markers of unidentified true disease susceptibility loci not present in linkage disequilibrium with the markers in East Africa ^(46)^. Therefore, *NRAMP1*-INT4 polymorphism may, in fact, serve as marker for other genes with a functional impact in ethnic-specific genetic backgrounds. Moreover, the unknown cofactors (such as socio-economic factors, nutritional status, or other co-infections) and the complex interactions between gene and other host factors as well as environmental factors might introduce differences and emphasize the difficulties to compare between studies.

Sudan has been estimated to have the highest incidence of *M. tuberculosis* infection in the Eastern Mediterranean region ^(15)^. From this view, more insight into the demographic characteristic of the TB patients and exploring the risk factors associated with TB is critical to provide a useful estimate of the potential for development of TB infections then improvement more effective preventive strategies. In this study, TB infection was more detected among males. Similar observation was noticed in other previous studies conducted on Sudanese population shown increased rate of TB infection among males ^(17, 18, 47, 48)^. The mean age of our patients is 32.7-year-old, many studies conducted in Sudan have reported the same results ^(18, 48, 49)^. However, these averages are considered lower when compared to the average mean above 50 which was noticed by other studies in the same area ^(50, 51)^.

Awareness about the importance of vaccination against TB infection seemed to be very confusing as the BCG scar failure rate was 96.7%. This indicates either those patients are not vaccinated, or the BCG scar disappears. BCG scar is a sensitive indicator of vaccination status up to one year after administration of the vaccine in the first month of life and the scar failure rate was 17% among Sudanese infants ^(52)^. The efficiency of BCG vaccine is very poor in our country. Gorish *et al*. showed that only 39% of previously healthy vaccinated adults were positive for Manteaux test, while 61% were negative and failed to develop induration post tuberculin test, and explained that by either improperly vaccination or invalid vaccine was taken ^(53)^. In this study, tobacco smoking was the most reportable risk factor (17%), followed by diabetes (10%), alcohol abuse (2%) and corticosteroid therapy (1%). Consistent with our result, a study in Spain has shown that tobacco smoking has direct action on the immune system and with alcohol consumption, drug abuse and contact with TB patients considered to be a higher risk factors of pulmonary tuberculosis ^(54)^. Ninety-one percent of patients presented with pulmonary TB, while 9% had extra-pulmonary TB. This was similar to the findings of many studies in which most of patients have pulmonary TB infection. For instance; in Ghana 78.2% of patients had PTB, while 21.8% had EPTB ^(55)^,70% had PTB in Spain, while 30% had EPTB ^(54)^, in Europe 80.3% reported with PTB and 19.3% had EPTB ^(56)^. Tuberculosis of bones and joints (Pott disease) was the most common presentation in EPTB patients. In contrary to a study conducted among the same population that revealed TB lymphadenitis as the most reportable EPTB type ^(48)^.

Analysis of tuberculosis drug resistant indicated that, 20% of previously treated patients have been diagnosed as MDR-TB, which were similar to the prevalence reported in Sudanese studies, 20% ^(49)^ and 22% ^(17)^. Compared with other studies in the region, the prevalence of MDR among previously treated patients double estimate for Ethiopia to the East (12%) ^(57)^, Kampala to the South (13%) ^(58)^ and less than the prevalence reported from Egypt (38%) ^(57)^.

## Conclusion

In this study, we found an association between heterozygosity for *NRAMP1*-INT4 variant and resistance to TB infection. Thus, the *NRAMP1*-INT4 polymorphism may serve as marker of unidentified genetic factors that may play a critical role in host immunity to TB in the Sudanese population. Further studies with large sample size are recommended to determine population-specific genetic associations with TB susceptibility in order to guide TB therapy and prophylaxis in an population-specific manner.

## Data Availability

The data regarding NRAMP1-INT4 genotypes and alleles distributions among participants are available from the corresponding author on reasonable request.

## Abbreviations

TB: Tuberculosis
NRAMP1: Natural Resistant Associated Macrophage Protein 1
INT4: Intron4
PTB: Pulmonary tuberculosis
EPTB: Extra-pulmonary tuberculosis
HIV: Human immune-deficiency virus
NGOs: Non-governmental Organizations
DST: drug susceptibility testing
SNP: Single nucleotide polymorphism
MDR-TB: Multi-drug resistance tuberculosis
MRI: Magnetic resonance imaging
BCG: Bacillus Calmete-Guérin
ZN: Ziehl-Neelsen Stain
EDTA: Ethylene-Diaminetetraacetic acid
PCR: Polymerase chain reaction
RFLP: Restriction fragment length polymorphism
OD: Odds ration
SPSS: statistical package for social science

## Acknowledgments

The authors are grateful for the patients and healthy controls who volunteer to participate in the study. The authors would like to gratefully acknowledge the assistance of Ahmed Mohamed Mansour in collection of control samples.

## Availability of data and materials

The data regarding *NRAMP1*-INT4 genotypes and alleles distributions among participants are available from the corresponding author on reasonable request.

## Authors’ contributions

HHM conceived and supervised the methodology. SGGM, HHM and AE designed the experiments. SGGM, AE, MAS, FOM and LAM performed the experiments. SGGM, THM, AE and ABI analyzed the data. SGGM wrote the manuscript. ABI and AE edited the manuscript. HHM edited and revised the final manuscript. All authors read and approved the final manuscript.

## Consent for publication

Not applicable

## Ethics approval and consent to participate

This study was approved by University of Khartoum, Faculty of Medical Laboratory Sciences review board, and Research Ethics Committee of Abu-Anja Specialized TB Hospital. Informed consent was obtained from all participants before they enrolled in the study.

